# MedSDoH: A Rule-Based System for Extracting Social Determinants of Health from Multi-site EHRs Based on the OHNLP Framework

**DOI:** 10.64898/2026.04.27.26351699

**Authors:** Jaerong Ahn, Sunyang Fu, Daniel Palacios, Hyun-Hwan Jeong, Liwei Wang, Maria C Swartz, Mustafa Tosur, Maria Jose Redondo, Xizhi Wu, Zhiyi Yue, Alexandra Kakadiaris, Nan Wang, Zhehan Li, Ming Huang, Andrew Wen, Daniel Harris, Yanshan Wang, Min Ji Kwak, Zhandong Liu, Hongfang Liu

## Abstract

**Objective:** Social Determinants of Health (SDoH) are critical to patient care and population health. Despite their importance, SDoH information is frequently embedded within unstructured clinical text such as patient-reported information or social worker notes, which limits its use on clinical decision-making and resource allocation. Although transformer-based models represent the current state of the art, their scalability, computational requirements, and limited transparency pose barriers to large-scale multi-site clinical implementation. In this context, rule-based NLP systems remain valuable, particularly when explainability, reproducibility, and rapid customization are essential.

**Methods:** MedSDoH was developed within the Open Health Natural Language Processing (OHNLP) Framework using literature-derived SDoH resources, standardized domain definitions, and expert-curated rulesets. Large language models (LLMs) were used during development to assist with rule generation and lexicon expansion. Rules were iteratively refined against a gold-standard annotated corpus from two health systems and then evaluated on independent datasets.

**Result:** The final system included 942 regular expression rules spanning 22 SDoH domains. On validation on two external datasets, MedSDoH demonstrated generalizability and comparable performance across sites. The system has been made publicly available so research community can collaboratively contribute to the maintenance and extension through disease- or site-specific adaptations.

**Conclusion:** MedSDoH is a computationally efficient and open-source system for large-scale SDoH extraction from clinical text. It is well-suited for multi-site adaptation and deployment in resource-constrained settings.

## Introduction

Social determinants of health (SDoH) encompass social, economic, and environmental conditions that influence health outcomes and population health, including factors such as income, education, housing, and access to resource.[1,2] Research has shown that improving SDoH can positively contribute to life expectancy, sanitation, gender equality, and primary school enrollment. [3–5] However, systematically capturing and utilizing SDoH information from electronic health records (EHRs) remains a challenge and it limits their integration into patient care and population health management.

One barrier lies in the nature of SDoH documentation itself. Despite their importance, SDoH factors such as housing instability, food insecurity, employment, transportation access, and social support are often underrepresented in structured EHR fields. For example, social history details are predominantly recorded in clinical text, rather than discrete, structured data fields.[6] While several initiatives have tried to capture SDoH information structurally, it has not substantially improved capture rates, with many institutions still relying on clinical text.[7–9] Extracting SDoH information from such text requires scalable, accurate, and interpretable computational methods.

Recent advancement in artificial intelligence, especially natural language processing (NLP), has enabled efficient extraction and standardization of SDoH information from clinical text.[10] Existing SDoH NLP-based solutions have employed methodologies ranging from rule-based, lexicon-based systems to machine learning models and transformer-based architecture.[11–13]These models face several practical implementation challenges. First, they are often difficult to generalize across institutions due to heterogeneity in clinical documentation. Second, training and deploying large models require substantial computational resources, making them inaccessible to many low-resource health settings. Third, most are black-box models that cannot be easily customized or iteratively improved by end users.

Many clinical outcomes are influenced by a multiple SDoH factors. [14] However, majority of existing NLP-empowered studies focused on extracting single, isolated SDoH variables, such as marital status, [15] occupation, [16] substance use, [12] residential instability, [17] and housing stability. [13] Moreover, significant heterogeneity exists in SDoH definitions, extraction methodologies, and validation approaches across studies, which limits the development of interoperable solutions. Consequently, the field lacks scalable, adaptable solutions that are transparent, and easily shared across diverse healthcare environments.

Here, we developed MedSDoH, a lightweight rule-based NLP system for extracting SDoH information from clinical text based on the Open Health Natural Language Processing (OHNLP) framework. The system was built through systematically synthesizing and standardizing SDoH resources curated from published SDoH literature. Relevant regex patterns and lexicons were generated with the assistance of leveraging large language models (LLMs). MedSDoH was iteratively refined by medical experts using a gold standard dataset and the finalized system was evaluated on datasets from two additional sites. The system was further validated with a heart failure cohort to demonstrate its real-world applicability. We demonstrate that MedSDoH provides accurate and efficient SDoH extraction solutions suitable for institutions with varying computational resources and infrastructure constraints. Our approach enables direct user-system interaction and local customization, offering high translational value across diverse healthcare settings.

## Methods

### Data Source

Social worker notes (n=1,990) were retrieved from patient cohorts diagnosed with type 1 diabetes (n=4,193) and type 2 diabetes (n=60,681) from the UT Physicians EHR systems using PheKB algorithms (https://www.phekb.org). For model development, 247 notes (97 from UT Physicians and 150 from Texas Children’s Hospital (TCH)) were annotated and used for SDoH rule generation and iterative refinement. For performance validation, 1,505 independently annotated notes (100 from TCH and 1,405 from the Medical Information Mart for Intensive Care (MIMIC)-IV dataset) were used.

### System Architecture

The MedSDoH system was developed and deployed within the Open Health Natural Language Processing (OHNLP) Framework to leverage its modular, interoperable, and extensible architecture for clinical NLP. The OHNLP framework provides a unified infrastructure that supports component-based processing pipelines, interoperable with HIT standards, and transparent workflow documentation, all critical for explainable and reproducible SDoH extraction. The OHNLP framework adopts the translational AI principle which puts real-world researchers and clinicians at the center of the process by allowing them to directly interact with the system and improve and customize it as local documentation patterns and priorities evolve. [18,19]

The rule-based extraction module of the OHNLP framework applies expert rules based on regular expressions, contextual rules, and dependency-based patterns to capture clinical concepts from text. The system applies rules to determine negation, temporality, and experiencer (patient vs. family). The extracted information is then standardized for downstream analysis.

### System Development and Evaluation

The system development and evaluation includes four tasks (Figure 1): (1) Ontology-guided SDoH concept definitions [20] via literature review and clinical document extraction from cohorts; (2) keyword annotation using the MedTator platform with iterative guideline development and multi-phase adjudication; (3) model development with LLM-assisted rule generation using GPT-4o to create rule-based NLP components, followed by iterative evaluation and fine-tuning; and (4) multi-site model evaluation across independent datasets with comprehensive statistical analysis. Each step is explained in detail below.

**Figure 1.**
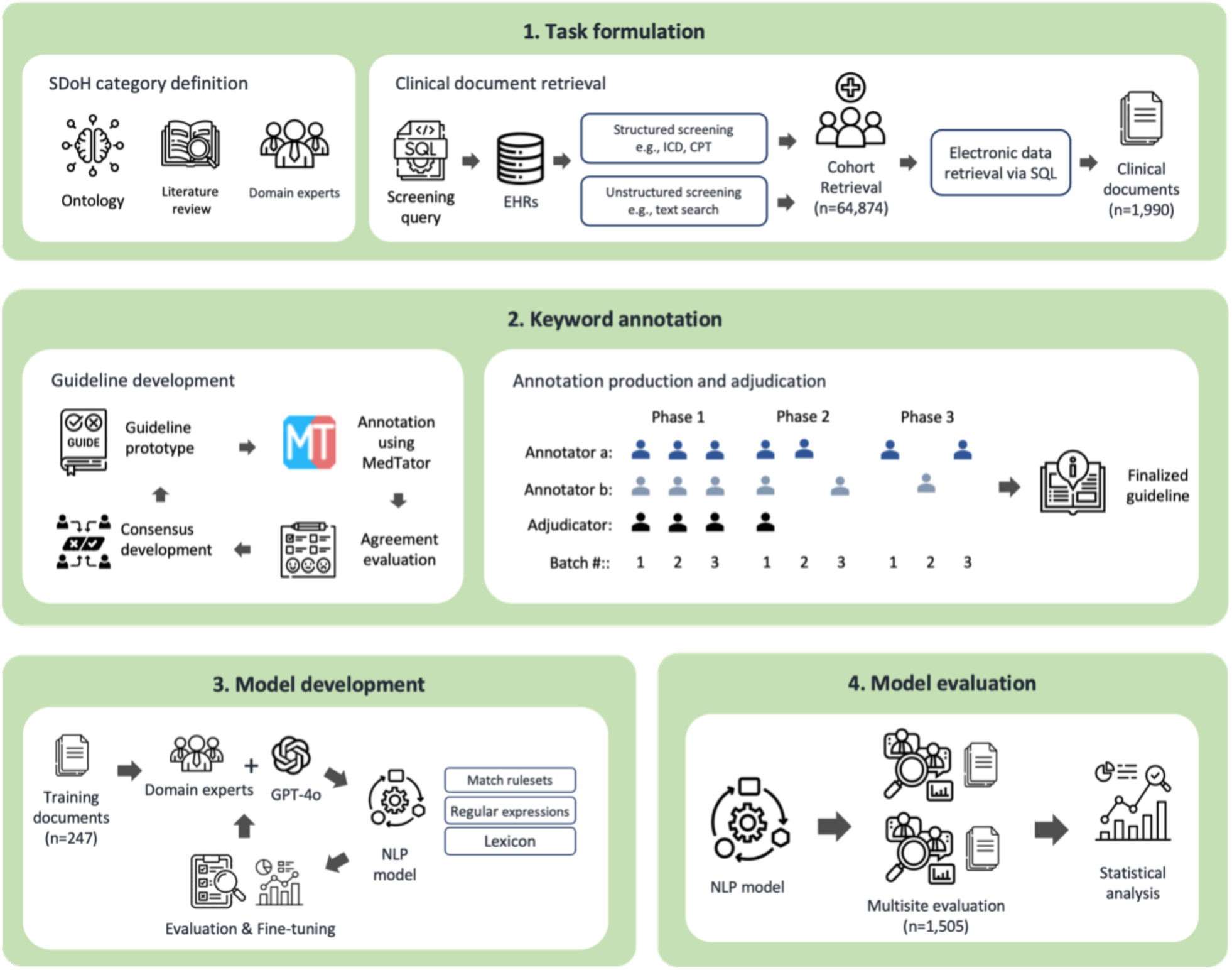
Schematic overview of the MedSDoH model development and validation workflows.

### SDoH Category definition and clinical document retrieval

For the current task, 67 peer-reviewed research articles with SDoH as a listed search keyword were collected for the study. Based on systematic literature review and established SDoH ontologies, [20] we defined the following 22 SDoH concept domains (**Appendix**).

### SDoH keyword annotation

Two expert annotators with clinical informatics background were recruited to manually label SDoH-related keywords on the social worker notes using MedTator, an open-source web-based annotation platform[21] (https://medtator.ohnlp.org). The annotation proceeded in multiple phases, beginning with guideline development, where clinical experts and NLP specialists collaboratively defined the annotation criteria for each SDoH category (Figure 1). Each annotator independently labeled an initial set of 20 notes, and the results were reviewed through an iterative adjudication process to refine the annotation guidelines and rule sets. Inter-annotator agreement (IAA) was calculated to assess and ensure consistency in the annotation results. The rulesets were refined until a sufficient IAA score of 0.907 was reached.

### Rule development

We employed a “human-in-the-loop” approach using an LLM (GPT-4o) as an assistant for generating regex patterns, identifying edge cases, and refining lexicons tailored to each SDoH domain. Once sufficient agreement was reached between the annotators, additional social worker notes (n=97 from UT Physicians and n=150 from Texas Children’s Hospital (TCH)) were randomly selected and labeled. These annotations served as the gold standard for evaluating the performance of the model-generated labels. Discrepancies between the model outputs and human annotations were systematically reviewed to optimize the rules. LLMs suggested potential expressions and keywords, which experts then validated or rejected before implementation. The refinement proceeded until the model achieved a high F1-score (∼0.9) on the training dataset.

### Multi-Site model evaluation

To assess the generalizability of the model, evaluation was conducted on 100 social worker notes from TCH and additional notes (n=1,405) obtained from the (MIMIC)-IV dataset.[22] The model outputs were compared with human annotations for each dataset. Key performance metrics, including precision, recall, and F1 score, as well as their macro-averages, were calculated. This multi-site validation approach ensured that the model remained robust across diverse clinical contexts and documentation formats.

### Feasibility demonstration on chronic heart failure patients

The feasibility of the current SDoH extraction framework was further tested on patients diagnosed with heart failure. A retrospective cohort (n=2,551) was established comprising patients aged 65 years and older who had at least two heart failure-related visits within a one-year period at UT Physicians’ internal medicine, family medicine, geriatric medicine, or cardiology clinics between January 1, 2021, and December 31, 2023. Using the MedSDOH framework, we extracted SDoH variables from 22 domains and employed chi-square tests to examine their associations with acute exacerbation events. This demonstration served as a real-world use case to evaluate the system’s potential to aid in clinical decision support. All statistical analyses were conducted using R (version 4.4.1).

## Results

### Multi-Site validation results

The MedSDoH system included 942 regex rules from 22 SDoH categories, with an average of 42.9 patterns per category (range: 6-165). The performance was validated using two independent datasets: MIMIC-IV and those obtained from TCH. The MedSDoH demonstrated consistent performance across both datasets, with F1-scores of 0.833 and 0.816 (precision: 0.768, recall: 1.000) in the TCH and MIMIC-IV datasets, respectively (Figure 2, Table 1). MIMIC-IV achieved perfect recall (1.000), but relatively lower precision (0.768), while TCH performance seemed more balanced (precision: 0.812, recall: 0.920) (Figure 2). The number of detected SDoH instances varied considerably, ranging from 1 to 41 in TCH, and 1 to 20 in MIMIC-IV (Table 1). Overall, the validation performance was comparable to the training performance obtained from our development sites, indicating that the model is generalizable across different EHR systems and institutional documentation practices.

**Figure 2.**
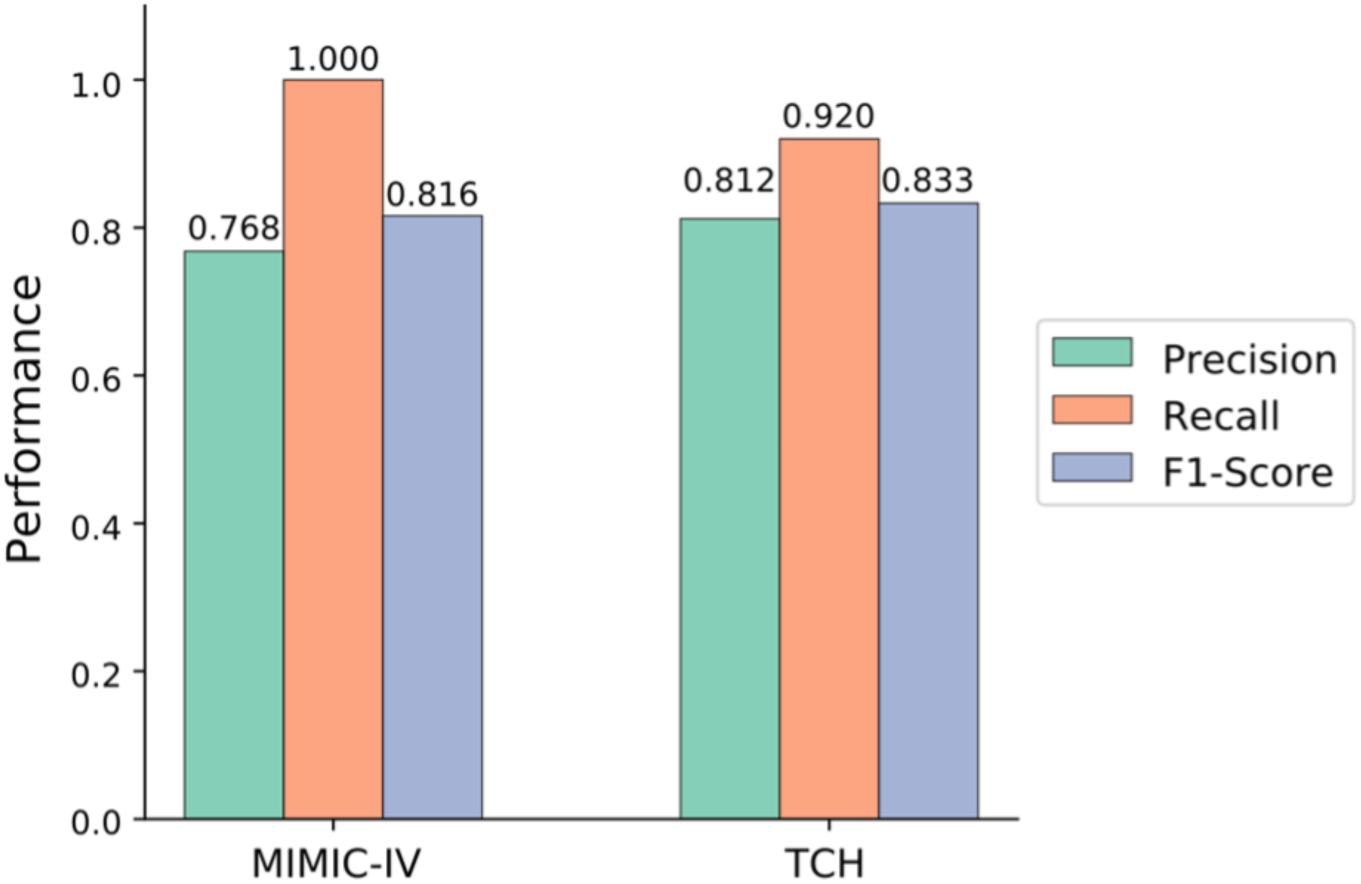
Multi-site validation performance results.

**Table 1.**
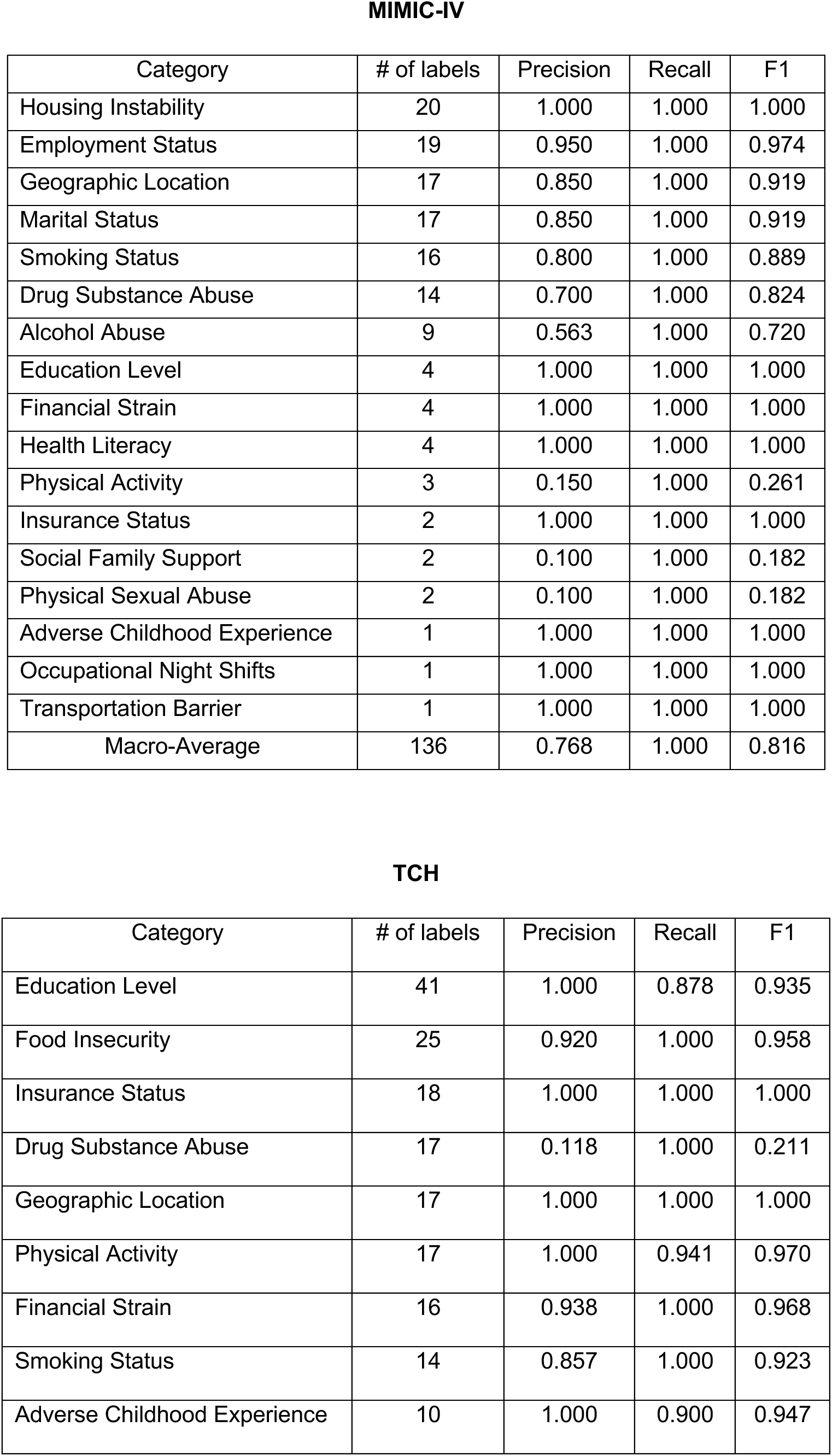
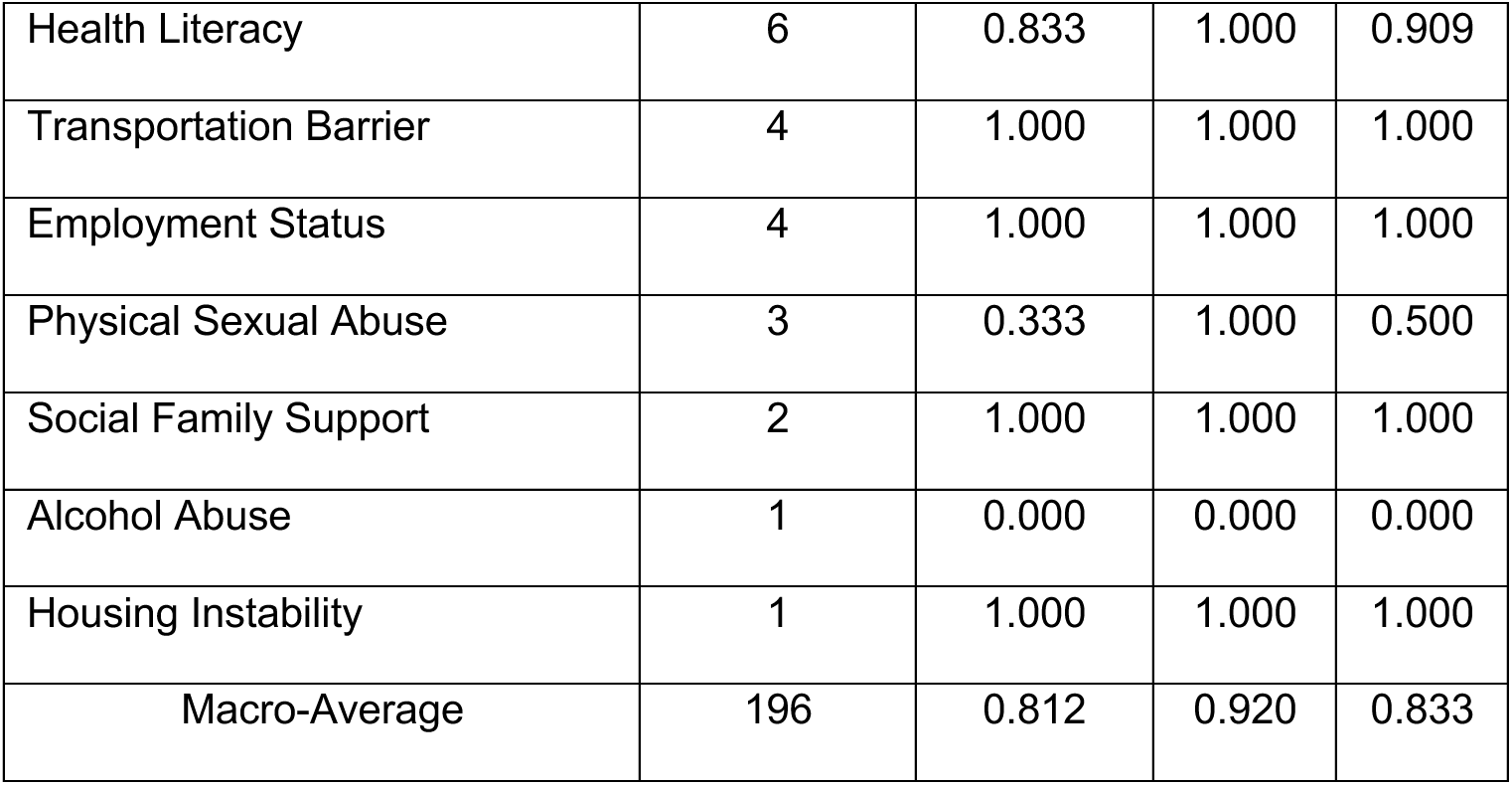
Multi-site validation results organized per SDoH category. Tables were sorted by the number of labels.

**MIMIC-IV**
| Category | # of labels | Precision | Recall | F1 |
| --- | --- | --- | --- | --- |
| Housing Instability | 20 | 1.000 | 1.000 | 1.000 |
| Employment Status | 19 | 0.950 | 1.000 | 0.974 |
| Geographic Location | 17 | 0.850 | 1.000 | 0.919 |
| Marital Status | 17 | 0.850 | 1.000 | 0.919 |
| Smoking Status | 16 | 0.800 | 1.000 | 0.889 |
| Drug Substance Abuse | 14 | 0.700 | 1.000 | 0.824 |
| Alcohol Abuse | 9 | 0.563 | 1.000 | 0.720 |
| Education Level | 4 | 1.000 | 1.000 | 1.000 |
| Financial Strain | 4 | 1.000 | 1.000 | 1.000 |
| Health Literacy | 4 | 1.000 | 1.000 | 1.000 |
| Physical Activity | 3 | 0.150 | 1.000 | 0.261 |
| Insurance Status | 2 | 1.000 | 1.000 | 1.000 |
| Social Family Support | 2 | 0.100 | 1.000 | 0.182 |
| Physical Sexual Abuse | 2 | 0.100 | 1.000 | 0.182 |
| Adverse Childhood Experience | 1 | 1.000 | 1.000 | 1.000 |
| Occupational Night Shifts | 1 | 1.000 | 1.000 | 1.000 |
| Transportation Barrier | 1 | 1.000 | 1.000 | 1.000 |
| Macro-Average | 136 | 0.768 | 1.000 | 0.816 |

| Category | # of labels | Precision | Recall | F1 |
| --- | --- | --- | --- | --- |
| Education Level | 41 | 1.000 | 0.878 | 0.935 |
| Food Insecurity | 25 | 0.920 | 1.000 | 0.958 |
| Insurance Status | 18 | 1.000 | 1.000 | 1.000 |
| Drug Substance Abuse | 17 | 0.118 | 1.000 | 0.211 |
| Geographic Location | 17 | 1.000 | 1.000 | 1.000 |
| Physical Activity | 17 | 1.000 | 0.941 | 0.970 |
| Financial Strain | 16 | 0.938 | 1.000 | 0.968 |
| Smoking Status | 14 | 0.857 | 1.000 | 0.923 |
| Adverse Childhood Experience | 10 | 1.000 | 0.900 | 0.947 |
| Health Literacy | 6 | 0.833 | 1.000 | 0.909 |
| Transportation Barrier | 4 | 1.000 | 1.000 | 1.000 |
| Employment Status | 4 | 1.000 | 1.000 | 1.000 |
| Physical Sexual Abuse | 3 | 0.333 | 1.000 | 0.500 |
| Social Family Support | 2 | 1.000 | 1.000 | 1.000 |
| Alcohol Abuse | 1 | 0.000 | 0.000 | 0.000 |
| Housing Instability | 1 | 1.000 | 1.000 | 1.000 |
| Macro-Average | 196 | 0.812 | 0.920 | 0.833 |

### Error Analysis

To better understand the model’s limitations, we conducted a comprehensive error analysis on a subset of labeling errors made by the model (n=201), following the established taxonomies for clinical NLP error analysis. [23] The identified errors spanned 14 SDoH categories such as Food Insecurity (n=47, 23.4%), Smoking Status (n=44, 21.9%), Social Family Support (n=33, 16.4%), etc. (Figure 3A). We identified 21 error types. Negation errors were the most common (n=52, 25.9%), followed by exclusion errors (n=49, 24.4%), together representing 50.2% of all errors (Figure 3B). This highlights the model’s primary challenge of handling negated statements. The result also aligns with previous multi-site NLP studies, where negation and exclusion represent the most common error categories in various EHR systems. [23]

**Figure 3.**
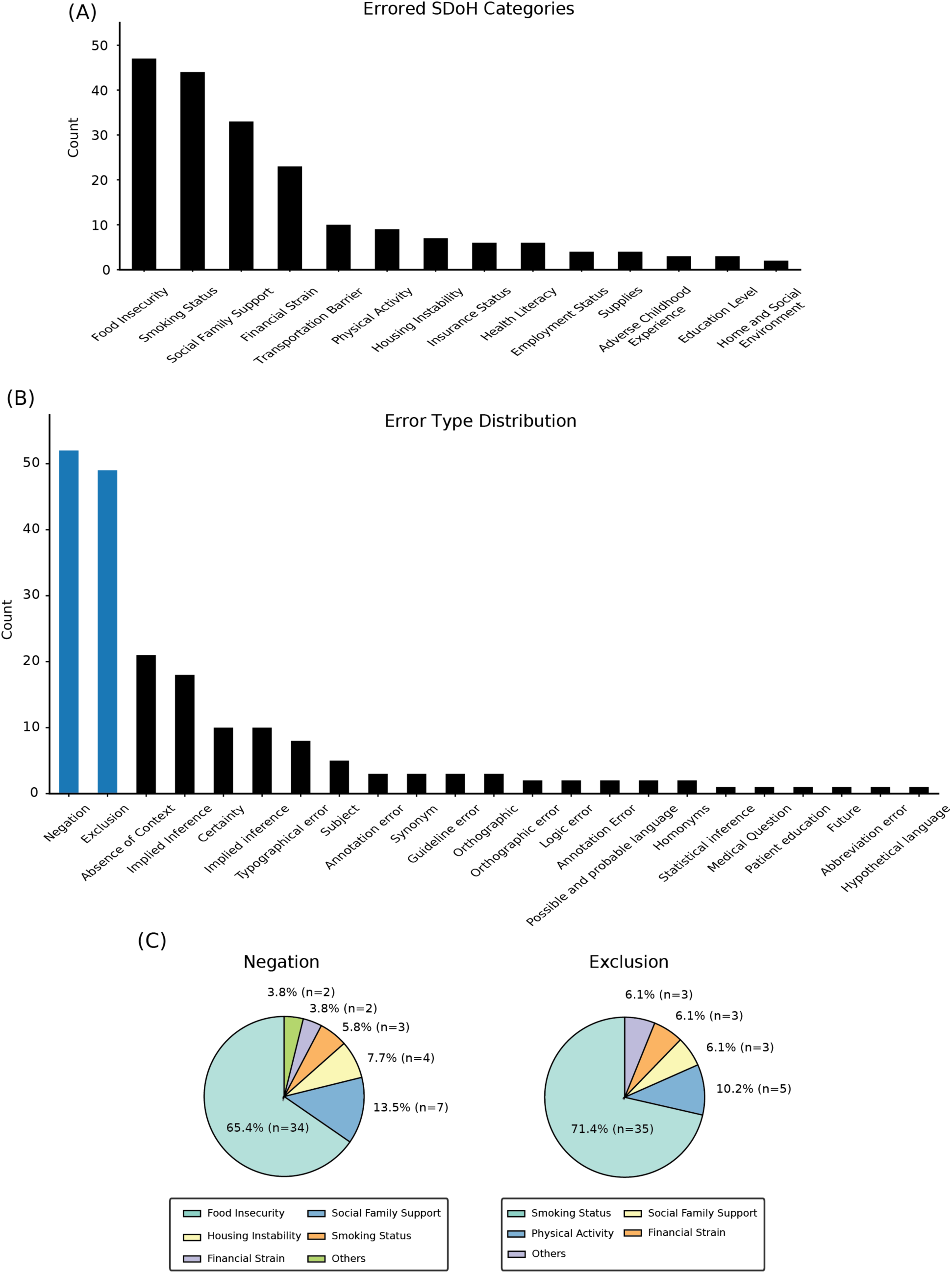
(A) Distribution of SDoH categories where errors were found. (B) Distribution of error types. Negation (n=52) and exclusion (n=49) errors (blue bars) accounted for 50.2% of all errors. (C) The two most prevalent error types (negation and exclusion) were broken down by SDoH categories.

The high frequency of negation errors primarily stems from the difficulty in correctly processing template-based documentation patterns that are commonly used for screening in clinical notes. For example, routine questionnaire phrases such as “food insecurity: no food insecurity” were frequently misclassified as positive SDoH mentions. These patterns are particularly challenging to capture because templates vary significantly across different EHR systems, making it difficult to develop universal negation rules that can be consistently applied. When the two most common errors were broken down by SDoH category, Food Insecurity was most dominant in negation errors (n=34) while Smoking Status was most prevalent in exclusion errors (n=35) (Figure 3C).

The next most common errors were absence of context (n=21) and implied inference (n=18) (Figure 3B). The prevalence of these errors reflects the fundamental limitations of rule-based models, which are particularly prone to failure when target information is expressed across multiple sentences or requires broader document-level understanding rather than keyword matching. Addressing these limitations will be an important future research direction.

### SDoH extraction from the patient cohort with heart failure

Among the identified cohort with chronic heart failure (n=2,551, see **Methods**), 673 (26%) had acute heart failure exacerbation (AHF) within 1 year. We assessed the association between AHF and SDoH factors using the Chi-square test. Among 22 SDoH factors the following yielded significant associations with AHF: financial strain (34% in AHF group vs 25% in non-AHF group, *p*-value <0.001), health literacy (24% in AHF group vs 28% in non-AHF group, *p*-value=0.029), physical activity (80% in AHF group vs 84% in non-AHF group, *p*-value= 0.028), supplies (2% in AHF group vs 0.5% in non-AHF group, *p*-value= 0.02). These preliminary findings demonstrate the system’s capacity to efficiently extract SDoH information at scale and the potential for downstream clinical research or decision support applications.

## Discussion

This study presents MedSDoH, a comprehensive rule-based NLP framework that addresses critical gaps in SDoH extraction from clinical documentation. Our approach demonstrates three key contributions: 1) the development of a standardized 22 category SDoH domains informed by published literature and established ontologies; 2) successful multi-site validation with robust performance across diverse EHR systems; 3) real-world clinical utility showing statistically significant associations between specific SDoH factors and heart failure exacerbations. These key findings establish our framework as a practical, scalable solution for institutions looking to integrate SDoH assessment into clinical workflows without substantial computational infrastructure.

### Comparison with previous research

Previous SDoH NLP studies have taken diverse methodological approaches, ranging from rule-based and lexicon-based systems to machine learning approaches using toolkits such as spaCy [24] and medspaCy.[25] Deep learning approaches primarily relied on transformer-based architectures such as BERT, RoBERTa, and BioClinicalBERT.[26–29] While deep learning models show promising performance, rule-based systems provide value for healthcare applications owing to their computational efficiency and minimal infrastructure requirements.

Also, most existing studies focused on singular SDoH concepts [12,13,15–17], with a handful of research tackling multiple SDoH categories. Those studies were comparatively limited in their scope, examining 7 social characteristics such as tobacco use, alcohol abuse, drug abuse, depression, housing instability, fall risk, and poor social support [30], and analyzing 9 life events including housing instability, job instability, justice involvement, social isolation, relationship issues, detoxification, military sexual trauma, access to lethal means, and food insecurity [31]. By contrast, our approach utilizes a standardized 22-category framework based on established ontologies [20] and comprehensive literature synthesis. Each category offers clear operational criteria that other researchers and institutions can adopt. This standardization addresses a current gap in the field, as many existing SDoH NLP frameworks are close-sourced, which limits comparative assessment.

### Computational efficiency and practical considerations

Current deep learning solutions also face critical barriers to large-scale clinical implementation. Although model-based approaches using transformers achieved high performance on specific tasks, they generally require substantial computational resources, which make them cost-prohibitive for processing millions of patients.[32] These models often lack the transparency and interpretability essential for healthcare applications.

Our rule-based system offers practical advantages for real-world clinical deployment. Once rulesets were developed with the assistance of LLMs, the system proceeds purely via pattern matching algorithms without any ongoing LLM costs. The system thus can handle large patient populations using standard hardware (even on a personal laptop or desktop). This contrasts with other proprietary cloud-based models like GPT variants, which can be prohibitively expensive and time-consuming when processing millions of patient records. These models, despite potential performance gains, require substantial investments with token processing costs that can exceed $100,000 for a single institutional dataset.[33]

Also, its transparent rule structure allows clinicians and informaticists to understand, validate, and modify the extraction logic directly. The rules can be customized for institution-specific documentation patterns and terminology. This level of customization would require extensive retraining with deep learning models.

### Limitations

Despite its advantages, our rule-based approach has inherent limitations that warrants discussion. Error analysis revealed that negation errors were most prominent (n=52 of 216 total errors), reflecting challenges in processing template-based documentation patterns. Institution-specific templates such as “food insecurity: no food insecurity” were frequently misclassified as positive SDoH mentions, demonstrating the difficulty of developing universal rules that function consistently across diverse EHR systems. The system also had difficulty in capturing SDoH when the information is expressed across multiple sentences or requires broader document-level understanding, as illustrated by other common error categories, such as absence of context (n=21) and implied inference (n=18). These limitations reflect trade-offs in prioritizing computational efficiency and interpretability over sophisticated contextual understanding.

It should be acknowledged that the current validation was conducted at two U.S. academic medical centers. Performance varied considerably across SDoH categories (Table 1), reflecting differences in documentation practices and template patterns. Users should interpret results with this variability in mind and may need to adjust performance thresholds for their specific applications. Further validation is needed across different hospital systems and clinical note types. We hope the open-source nature of MedSDoH will gain broader adoption in the research community and facilitate such testing across diverse healthcare settings.

Additionally, our current approach identifies only the presence of SDoH mentions without capturing the direction or magnitude of their impact, making it difficult to pinpoint whether an SDoH factor represents a positive resource or negative burden for patient outcomes. Future studies will aim to address these limitations via directional modeling.

### Future directions

Several research directions could enhance MedSDoH while preserving its core advantages. Enhanced negation detection through more sophisticated template recognition could improve its precision. An additional evaluation layer powered by a fine-tuned LLM could potentially correct rule outputs and recursively refine regex patterns, combining rule efficiency with improved contextual understanding.

The system showed promise in generalizing beyond the diabetes cohorts used for initial development. The observed associations between specific SDoH factors and heart failure exacerbations, while preliminary, demonstrate the feasibility of combining automated SDoH extraction with clinical outcomes data. Future study with multivariable models adjusting potential confounders (e.g., age and comorbidities) to rigorously examine the predictive potential of this framework for clinical decision support is needed. The lightweight architecture makes the system well-suited for integration with real-time clinical decision support systems, potentially enabling identification of high-risk patients for targeted interventions.

## Data Availability

The complete MedSDoH framework, including SDoH category definitions, rule sets, source code, and documentation, is available at https://github.com/OHNLP/MedSDoH The institutional EHR data and annotated clinical notes used in the study contain protected health information and are not publicly available. Access may be considered by the contributing institutions subject to institutional review board approval and applicable data use agreements.

## Author Contributions

J.A. (Conceptualization, Data curation, Formal analysis, Investigation, Methodology, Software, Validation, Visualization, Writing – original draft, Writing – review & editing); S.F. (Conceptualization, Data curation, Formal analysis, Funding acquisition, Investigation, Methodology, Project administration, Resources, Software, Supervision, Writing – original draft, Writing – review & editing); D.P. (Data curation, Formal analysis, Validation, Writing – review & editing); H.H.J. (Formal analysis, Software, Validation, Writing – review & editing); L.W. (Data curation, Project administration, Supervision, Validation); M.C.S. (Funding acquisition, Project administration, Validation, Writing – review & editing); M.T. (Data curation, Project administration, Writing – review & editing); M.J.R. (Resources, Supervision, Writing – review & editing); X.W. (Data curation, Formal analysis); Z.Y. (Validation); A.K. (Validation); N.W. (Validation); Z.L. (Data curation, Validation); M.H. (Data curation, Resources); A.W. (Resources, Software); D.R.H. (Writing – review & editing); Y.W. (Writing – review & editing); M.J.K. (Supervision, Writing – review & editing); Z.L.Z. (Project administration, Supervision, Writing – review & editing); H.L. (Conceptualization, Funding acquisition, Investigation, Project administration, Supervision, Writing – original draft, Writing – review & editing)

## References

1 Braveman P, Gottlieb L. The Social Determinants of Health: It’s Time to Consider the Causes of the Causes. Public Heal Rep. 2014;129:19–31. doi: 10.1177/00333549141291s206

2 Daniel H, Bornstein SS, Kane GC, et al. Addressing Social Determinants to Improve Patient Care and Promote Health Equity: An American College of Physicians Position Paper. Ann Intern Med. 2018;168:577–8. doi: 10.7326/m17-2441

3 Calley DQ, Fu S, Hamilton MD, et al. Assessment of Gender Differences in Letters of Recommendation for Physical Therapy Residency Applications. J Phys Ther Educ. 2024;38:331–9. doi: 10.1097/jte.0000000000000337

4 Fu S, Calley DQ, Rasmussen VA, et al. Gender-based Language Differences in Letters of Recommendation. AMIA Jt Summits Transl Sci Proc AMIA Jt Summits Transl Sci. 2023;2023:196–205.

5 Hauck K, Martin S, Smith PC. Priorities for action on the social determinants of health: Empirical evidence on the strongest associations with life expectancy in 54 low-income countries, 1990–2012. Soc Sci Med. 2016;167:88–98. doi: 10.1016/j.socscimed.2016.08.035

6 Chen ES, Manaktala S, Sarkar IN, et al. A multi-site content analysis of social history information in clinical notes. AMIA Annu Symp Proc AMIA Symp. 2011;2011:227–36.

7 Gold R, Bunce A, Cowburn S, et al. Adoption of Social Determinants of Health EHR Tools by Community Health Centers. Ann Fam Med. 2018;16:399–407. doi: 10.1370/afm.2275

8 Cohen DJ, Wyte-Lake T, Dorr DA, et al. Unmet information needs of clinical teams delivering care to complex patients and design strategies to address those needs. J Am Méd Inform Assoc. 2020;27:690–9. doi: 10.1093/jamia/ocaa010

9 Wang M, Pantell MS, Gottlieb LM, et al. Documentation and review of social determinants of health data in the EHR: measures and associated insights. J Am Méd Inform Assoc. 2021;28:2608–16. doi: 10.1093/jamia/ocab194

10 Bejan CA, Angiolillo J, Conway D, et al. Mining 100 million notes to find homelessness and adverse childhood experiences: 2 case studies of rare and severe social determinants of health in electronic health records. J Am Méd Inform Assoc. 2018;25:61–71. doi: 10.1093/jamia/ocx059

11 Han S, Zhang RF, Shi L, et al. Classifying social determinants of health from unstructured electronic health records using deep learning-based natural language processing. J Biomed Inform. 2022;127:103984. doi: 10.1016/j.jbi.2021.103984

12 Afshar M, Phillips A, Karnik N, et al. Natural language processing and machine learning to identify alcohol misuse from the electronic health record in trauma patients: development and internal validation. J Am Méd Inform Assoc. 2019;26:254–61. doi: 10.1093/jamia/ocy166

13 Chapman AB, Jones A, Kelley AT, et al. ReHouSED: A novel measurement of Veteran housing stability using natural language processing. J Biomed Inform. 2021;122:103903. doi: 10.1016/j.jbi.2021.103903

14 Kangas T, Milis S-L, Vanthomme K, et al. The social determinants of health-related quality of life among people with chronic disease: a systematic literature review. Qual Life Res. 2025;34:2501–11. doi: 10.1007/s11136-025-03976-1

15 Bucher BT, Shi J, Pettit RJ, et al. Determination of Marital Status of Patients from Structured and Unstructured Electronic Healthcare Data. AMIA Annu Symp Proc AMIA Symp. 2019;2019:267–74.

16 Chilman N, Song X, Roberts A, et al. Text mining occupations from the mental health electronic health record: a natural language processing approach using records from the Clinical Record Interactive Search (CRIS) platform in south London, UK. BMJ Open. 2021;11:e042274. doi: 10.1136/bmjopen-2020-042274

17 Hatef E, Rouhizadeh M, Nau C, et al. Development and assessment of a natural language processing model to identify residential instability in electronic health records’ unstructured data: a comparison of 3 integrated healthcare delivery systems. JAMIA Open. 2022;5:ooac006. doi: 10.1093/jamiaopen/ooac006

18 Fu S, Wang L, Moon S, et al. Recommended practices and ethical considerations for natural language processing-assisted observational research: A scoping review. Clin Transl Sci. 2023;16:398–411. doi: 10.1111/cts.13463

19 Liu S, Wen A, Wang L, et al. An open natural language processing (NLP) framework for EHR-based clinical research: a case demonstration using the National COVID Cohort Collaborative (N3C). J Am Méd Inform Assoc. 2023;30:2036–40. doi: 10.1093/jamia/ocad134

20 Dang Y, Li F, Hu X, et al. Systematic design and data-driven evaluation of social determinants of health ontology (SDoHO). J Am Méd Inform Assoc. 2023;30:1465–73. doi: 10.1093/jamia/ocad096

21 He H, Fu S, Wang L, et al. MedTator: a serverless annotation tool for corpus development. Bioinformatics. 2022;38:1776–8. doi: 10.1093/bioinformatics/btab880

22 Johnson AEW, Bulgarelli L, Shen L, et al. MIMIC-IV, a freely accessible electronic health record dataset. Sci Data. 2023;10:1. doi: 10.1038/s41597-022-01899-x

23 Fu S, Wang L, He H, et al. A taxonomy for advancing systematic error analysis in multi-site electronic health record-based clinical concept extraction. J Am Méd Inform Assoc. 2024;31:1493–502. doi: 10.1093/jamia/ocae101

24 Mitra A, Ahsan H, Li W, et al. Risk Factors Associated With Nonfatal Opioid Overdose Leading to Intensive Care Unit Admission: A Cross-sectional Study. JMIR Méd Inform. 2021;9:e32851. doi: 10.2196/32851

25 Lybarger K, Dobbins NJ, Long R, et al. Leveraging natural language processing to augment structured social determinants of health data in the electronic health record. J Am Méd Inform Assoc. 2023;30:1389–97. doi: 10.1093/jamia/ocad073

26 Luo Y, Thompson WK, Herr TM, et al. Natural Language Processing for EHR-Based Pharmacovigilance: A Structured Review. Drug Saf. 2017;40:1075–89. doi: 10.1007/s40264-017-0558-6

27 Alsentzer E, Murphy JR, Boag W, et al. Publicly Available Clinical BERT Embeddings. arXiv. Published Online First: 2019. doi: 10.48550/arxiv.1904.03323

28 Wu S, Roberts K, Datta S, et al. Deep learning in clinical natural language processing: a methodical review. J Am Méd Inform Assoc. 2020;27:457–70. doi: 10.1093/jamia/ocz200

29 Yu Z, Yang X, Dang C, et al. A Study of Social and Behavioral Determinants of Health in Lung Cancer Patients Using Transformers-based Natural Language Processing Models. AMIA Annu Symp Proc AMIA Symp. 2022;2021:1225–33.

30 Navathe AS, Zhong F, Lei VJ, et al. Hospital Readmission and Social Risk Factors Identified from Physician Notes. Heal Serv Res. 2018;53:1110–36. doi: 10.1111/1475-6773.12670

31 Morrow D, Zamora-Resendiz R, Beckham JC, et al. A case for developing domain-specific vocabularies for extracting suicide factors from healthcare notes. J Psychiatr Res. 2022;151:328–38. doi: 10.1016/j.jpsychires.2022.04.009

32 Li H, Fu J-F, Python A. Implementing Large Language Models in Health Care: Clinician-Focused Review With Interactive Guideline. J Méd Internet Res. 2025;27:e71916. doi: 10.2196/71916

33 Fu S, Kwak MJ, Ahn J, et al. Advancing Delirium Detection through the Open Health Natural Language Processing Consortium and ENACT Network. J Gerontol, Ser A: Biol Sci Méd Sci. 2025;glaf207. doi: 10.1093/gerona/glaf207

