## Appendix for "MedSDoH: A Rule-Based System for Extracting Social Determinants of Health from Multi-site EHRs Based on the OHNLP Framework"

| SDoH Category | Definition | Impact | Interpretation |
| --- | --- | --- | --- |
| Adverse Childhood Experience | Adverse childhood experiences, or ACEs, are potentially traumatic events that occur in childhood (0-17 years). Examples include experiencing violence, abuse, or neglect; witnessing violence in the home or community; having a family member attempt or die by suicide. Also included are aspects of the child's environment that can undermine their sense of safety, stability, and bonding, such as growing up in a household with substance use problems, mental health problems, instability due to parental separation, or household members in jail or prison. | negative | Presence indicates the patient experienced adverse childhood events. |
| Alcohol Abuse | Alcohol use disorder (AUD) is a medical condition characterized by an impaired ability to stop or control alcohol use despite adverse social, occupational, or health consequences­. | negative | Presence indicates the patient is experiencing or has experienced alcohol abuse. |
| Drug Substance Abuse | The use of illegal drugs or the use of prescription or over-the-counter drugs for purposes other than those for which they are meant to be used, or in excessive amounts. Drug abuse may lead to social, physical, emotional, and job-related problems. | negative | Presence indicates the patient is experiencing or has experienced drug or substance abuse. |
| Education Level | Educational attainment categories including: no formal education, some high school, high school graduate, some college, associate degree graduate, college (4-year) graduate, some graduate school, graduate school graduate (MS, MA), and graduate school graduate (PhD, JD, MD). | N/A | Captures patient's education level. |
| Employment Status | Employment categories including unemployed, retired, on disability, homemaker, and other/uncertain status. | negative | Presence indicates the patient is unemployed or not in the workforce. |
| Environmental Exposure | Contact with chemical, biological, or physical substances found in air, water, food, or soil that may have a harmful effect on a person's health. | negative | Presence indicates harmful environmental exposure. |
| Financial Strain | Lack of cash or savings, or long-term or short-term loss of reliable income sources. Evidence of requiring assistance to afford living basics. | negative | Presence indicates the patient is experiencing or has experienced financial strain. |
| Food Insecurity | Unable to access nutritious and affordable food. Evidence of requiring assistance to secure food supply. | negative | Presence indicates the patient is experiencing or has experienced food insecurity. |
| Gender Identity | The personal sense of one's own gender. Gender identity can correlate with a person's assigned sex or can differ from it. In most individuals, the various biological determinants of sex are congruent and consistent with the individual's gender identity. | N/A | Captures patient's gender identity. |
| Geographic Location | Information about location born and location raised. | N/A | Captures patient's geographic background. |
| Health Literacy | Gap in administrative health literacy, such as English language, insurance, or medical assistance application difficulties. | negative | Presence indicates the patient has low health literacy. |
| Home and Social Environment | Lack of safety, insecurity, or poor physical condition of one's home or social surroundings. | negative | Presence indicates the patient is living in an inadequate social environment. |
| Housing Instability | Instability, such as homelessness or difficulty paying rent or utilities. | negative | Presence indicates the patient is experiencing or has experienced housing instability. |
| Insurance Status | Health insurance contract that requires the health insurer to pay some or all health care costs in exchange for a premium. | negative | Presence indicates the patient lacks adequate health insurance. |
| Occupational Night Shift | Work schedules where the primary working hours occur during the nighttime. | negative | Presence indicates the patient works night shifts. |
| Physical Activity | Any bodily movement produced by skeletal muscles that requires energy expenditure, including movement during leisure time, for transport, or as part of work or domestic activities. Both moderate- and vigorous-intensity physical activity improve health. | N/A | Presence indicates the patient's engagement in physical activity; requires context to interpret. |
| Physical Sexual Abuse | Physical abuse is intentional bodily injury, including slapping, pinching, choking, kicking, shoving, or inappropriately using drugs or physical restraints. Sexual abuse is nonconsensual sexual contact, including unwanted touching, rape, sodomy, coerced nudity, or sexually explicit photographing. | negative | Presence indicates the patient is experiencing or has experienced physical or sexual abuse. |
| Smoking Status | Recoded variable based on cigarette smoking questions, including categories of current smoker, former smoker, never smoked, and smoking status unknown. Includes vaping. | negative | Presence indicates the patient smokes or has smoked. |
| Social Family Support | Lack of physical and emotional comfort derived from family, friends, colleagues, and community. Includes constructs such as emotional support, instrumental support, and social network, which are associated with improved health outcomes. Social isolation is associated with higher health care expenditure, morbidity, and mortality. | negative | Presence indicates the patient lacks social or family support. |
| Supplies | Lack of availability, stability, and adequacy of essential resources that support daily living and well-being. | negative | Presence indicates the patient lacks adequate supplies. |
| Technical Literacy | The ability to use, understand, and manage technology safely, effectively, and responsibly. Includes the ability to create, evaluate, and integrate information using technology. | negative | Presence indicates the patient lacks technical literacy. |
| Transportation Barrier | Transportation issues due to socioeconomic constraints that prevent access to goods and services, such as health care visits. | negative | Presence indicates the patient is experiencing or has experienced a transportation barrier. |
